# Correlates of time to presentation for stroke care among patients at a tertiary hospital in Ondo State, Nigeria: A retrospective records review

**DOI:** 10.64898/2026.06.06.26355064

**Authors:** Olubusayo Ogunsemoyin, Olufunke Fayehun

## Abstract

**Introduction:** Early hospital presentation after stroke onset is necessary for rapid assessment and access to time-dependent acute management. This study examined the correlates of late presentation for stroke care among patients recorded at a tertiary hospital in Ondo State, Nigeria.

**Methods:** A retrospective records review was conducted using secondary data from the Stroke Registry of the University of Medical Sciences Teaching Hospital, radiology department records, referral notes, and ambulance records. Records of stroke cases documented within the preceding 24 months were reviewed. Late presentation was defined as hospital presentation more than four hours after symptom onset. Frequencies, chi-square tests, and modified Poisson regression with robust standard errors were used to estimate adjusted prevalence ratios.

**Results:** The analysis included 371 stroke cases. Of these, 317 (85.4%) presented after four hours, and the median time to presentation was 24 hours (interquartile range: 9-72 hours). Late presentation differed significantly by employment status, first-contact route, and pathway complexity at bivariate analysis. After adjustment, non-hospital first contact remained strongly associated with late presentation: patients whose first documented contact was non-hospital-based had almost 3 times the prevalence of delay compared with those whose first contact was hospital-based (adjusted prevalence ratio = 2.89; 95% confidence interval: 2.15-3.90; p < 0.001).

**Conclusion:** Late presentation was pervasive in this tertiary hospital record cohort and was primarily associated with the initial direction of care-seeking. Stroke response interventions should emphasise immediate hospital presentation and strengthen urgent referral from non-hospital first-contact points.

## Introduction

Stroke is a time-dependent neurological emergency and a major public health problem. The 2025 World Stroke Organisation fact sheet and the Global Burden of Disease 2021 analysis indicate that stroke remains among the leading causes of death and disability worldwide, with rising absolute numbers of incident strokes, stroke-related deaths and disability-adjusted life-years since 1990 [1,2]. In 2021, stroke accounted for about 7.3 million deaths and 160.5 million disability-adjusted life-years globally, while 93.8 million people were living with stroke, and 11.9 million new strokes occurred [2]. This burden is not evenly distributed. Low- and middle-income countries bear a disproportionate share of stroke mortality, disability and service gaps, reflecting the interplay of epidemiological transition, population ageing, hypertension and other vascular risk factors, poverty, limited emergency transport and uneven access to organised stroke care [1-3].

Nigeria is part of this wider inequity. Stroke has long been a leading cause of adult neurological admissions, death and disability in the country. Yet, the services needed for rapid diagnosis and early management are not consistently available. African evidence shows high incidence and case fatality, with pooled estimates indicating substantial early mortality after stroke [4,5]. Reviews of African stroke care describe persistent shortages of emergency medical transport, stroke-trained personnel, stroke units, affordable brain imaging, thrombolysis and rehabilitation services [6-8]. In Nigeria specifically, a nationwide survey identified dedicated stroke units in only five of 58 tertiary hospitals. At the same time, a review of out-of-hospital emergency care found the emergency response system to be fragmented and unevenly developed [9,10]. These gaps matter because the value of acute stroke treatment depends on how quickly patients reach a facility that can assess, image, triage, and refer or treat them.

Time is central to the biological and clinical rationale for this study. Evidence underpinning acute ischaemic stroke care shows that the probability of functional recovery falls as onset-to-treatment time increases. The NINDS trial established the efficacy of intravenous thrombolysis within three hours of onset, and later trials and meta-analyses showed benefit for selected patients treated up to 4.5 hours, with earlier treatment producing larger proportional benefit [11-13]. The phrase ‘time is brain’ captures the rapid loss of salvageable neural tissue during untreated ischaemia [14]. Mechanical thrombectomy has extended treatment opportunities for selected patients with large-vessel occlusion, including imaging-selected patients in later windows, but these advances do not make delay harmless; they make timely recognition, imaging and referral even more important [15-18]. Current AHA/ASA and European guidance therefore emphasises rapid prehospital recognition, emergency medical system activation, urgent brain imaging and streamlined transfer to appropriate acute stroke services [19-21].

Late presentation is a persistent barrier to acute stroke care in Africa. A 2025 systematic review and meta-analysis estimated that approximately four in five stroke patients in African studies experienced prehospital delay; poor awareness of stroke symptoms and longer distances to health facilities were significantly associated factors [22]. Earlier African reviews similarly reported that only a minority of patients reached the hospital within the first few hours after symptom onset and highlighted weak ambulance systems and limited stroke service capacity as structural contributors to late arrival [6]. Nigerian evidence is consistent with this pattern. In Calabar, Philip-Ephraim and colleagues documented substantial prehospital delay and linked late arrival to poor recognition of stroke symptoms [23]. In Ibadan, Ogbole and colleagues reported a mean time to computed tomography imaging of 70 hours; only 31% presented within 12 hours, and none within 3 hours of symptom onset [24]. These findings indicate that late presentation is not simply an individual-level problem; it is produced within a broader ecology of health literacy, transport, referral, affordability and service readiness.

One weakness in the literature is that late presentation is often treated as reflecting only patient ignorance or indecision. That framing is too narrow for a pluralistic health system. In Nigeria and many other African settings, households may respond to sudden illness through home treatment, patent medicine vendors, private clinics, prayer houses, traditional healers, primary health centres, general hospitals, or sequential combinations of these options. Evidence from studies of health-care use in Nigerian contexts shows that care seeking is shaped by perceived severity, cost, proximity, trust, social networks, provider availability, and local interpretations of illness [25-27]. Stroke symptoms may therefore prompt prompt action, but not necessarily towards immediate access to emergency neuroimaging or an acute stroke-ready service.

Public awareness studies reinforce this concern. Nigerian studies have documented uneven knowledge of stroke warning signs, risk factors and appropriate emergency response, even among high-risk groups and educated populations [28-30]. A community-based study in Benin City found relatively good recognition of some risk factors and symptoms, but also showed that faith-based settings were a major source of stroke information. In contrast, hospital settings contributed much less [31]. Such findings are important because information channels shape what families define as urgent, where they go first, and how quickly they move from symptom recognition to hospital presentation. For acute stroke, the first action may be as important as recognition itself: recognising weakness, speech disturbance, or facial deviation does not improve treatment eligibility if the first response is home management, a medicine vendor, spiritual care or another non-hospital route that delays imaging and referral.

Existing Nigerian and African studies have established that delay is common and have identified factors such as symptom awareness, distance, transport, consultation patterns and stroke severity [6,8,22-24]. However, there remains a need for local evidence linking delay to the documented sequence of care-seeking before arrival at a tertiary hospital. This is particularly relevant in Ondo State, where tertiary stroke care is accessed through a mix of self-referral, referrals from formal facilities and care initiated outside hospital settings. Understanding the first point of contact is analytically important because it is a modifiable pathway feature: interventions can target households, community actors, patent medicine vendors, faith-based providers, traditional providers, primary care facilities and referral systems to shorten the route from symptom onset to hospital assessment.

The present study, therefore, examined the correlates of late presentation for stroke care among patients identified from secondary hospital records at the University of Medical Sciences Teaching Hospital in Ondo State, Nigeria. It problematises late presentation as a pathway issue rather than an awareness deficit alone. Specifically, it asked: how common was presentation more than 4 hours after symptom onset, and were sociodemographic characteristics, pathway complexity, and the first documented care-seeking route associated with late presentation? By focusing on the first-contact route, the study aims to generate evidence to inform context-sensitive stroke response interventions, with particular attention to immediate hospital-directed action and urgent referral from common non-hospital first-contact points.

### Theoretical framework

This study was guided by Andersen’s Behavioural Model of Health Services Use, which explains utilisation as a function of predisposing characteristics, enabling resources and need-related factors [32]. In the available secondary-record data, age, sex, marital status and ethnicity were treated as predisposing characteristics. In contrast, employment status and place of residence were treated as enabling or access-related characteristics. Because the outcome concerns timely entry into hospital-based stroke care, the first documented point of contact was treated as the principal utilisation variable.

A care-pathway lens was used alongside Andersen’s model. This lens highlights the sequence of decisions and contacts linking symptom onset to arrival at a tertiary hospital. In pluralistic health systems, formal biomedical care coexists with informal, commercial, spiritual and traditional sources of help [25-27]. A direct first contact with hospital services, therefore, differs from an initial home-based, medicine-vendor, traditional, spiritual or other non-hospital contact, as additional steps may be required before specialist evaluation. The analytic premise is that delay may arise not only from whether patients seek help, but also from where the pathway begins and the number of steps taken before hospital presentation.

## Materials and methods

### Study design and setting

This retrospective hospital record-based study was conducted at the University of Medical Sciences Teaching Hospital (UNIMEDTH), Ondo State, Nigeria. UNIMEDTH is a tertiary referral facility that provides specialist services, including the evaluation and management of patients presenting with suspected or documented stroke.

### Data sources and study population

Quantitative data were drawn from multiple secondary sources: the UNIMEDTH Stroke Registry, radiology department records, referral notes, and ambulance records. These records were used to identify stroke cases reported in the 24 months preceding the study and to obtain information relevant to the present analysis. No direct participant interviews or prospective follow-up were conducted for this paper. The data were accessed for research purposes from 22 July 2024 to 17 January 2025.

The analytic dataset comprised 371 recorded stroke cases with available information on age, sex, marital status, work status, ethnicity, place of residence, duration from symptom onset to tertiary hospital presentation, the recorded care-seeking pathway, and whether the first contact was hospital or non-hospital. Because the study relied on clinical and administrative records, the analysis was limited to variables consistently documented in the compiled dataset.

### Outcome variable

The primary outcome was late presentation for stroke care. The interval from symptom onset to presentation at UNIMEDTH was recorded in hours. For the primary analysis, cases were classified as early presentation if arrival occurred within 4 hours of symptom onset, and as late presentation if arrival occurred more than 4 hours after symptom onset. A sensitivity classification based on presentation after 4.5 hours was also examined, as this threshold corresponds to a commonly used treatment window in acute ischaemic stroke care. The classifications were identical in this dataset because no case was recorded between 4.0 and 4.5 hours.

### Explanatory variables

The explanatory variables were age group (<45, 45-64 and 65+ years), sex (male or female), marital status (married or not married), employment status (employed or unemployed), place of residence (urban or rural), ethnicity (Yoruba or non-Yoruba) and first-contact route (hospital or non-hospital). The main exposure of interest was the first-contact route. Hospital first contact included cases whose first documented contact was with a hospital or primary health centre. Non-hospital first contact included care initiated through self-medication, chemist/patent medicine use, traditional home remedies, traditional healers, prayer houses, or home management by a doctor or nurse before hospital presentation.

Pathway complexity was defined as the number of documented care-seeking steps before or at presentation at UNIMEDTH, ranging from 1 to 4. It was examined descriptively and bivariately. It was not included in the primary adjusted model because it may occur after the initial first-contact route and be partly generated by it.

### Statistical analysis

Frequencies and percentages were used to describe categorical variables. The median and interquartile range were used to summarise the symptom-onset-to-presentation interval because the distribution was markedly right-skewed. Associations between each explanatory variable and late presentation were initially assessed using Pearson chi-square tests; for two-by-two tables, the continuity correction was applied.

A modified Poisson regression with robust standard errors was fitted to estimate adjusted prevalence ratios (aPRs) and 95% confidence intervals for late presentation. This approach was chosen because late presentation was common in the analytic dataset and because prevalence ratios are more interpretable than odds ratios when the binary outcome is highly prevalent [33,34]. The adjusted model included age group, sex, marital status, employment status, residence, ethnicity and first-contact route. Estimates were reproduced in Python using the pandas, scipy and statsmodels packages. Statistical significance was assessed at p < 0.05.

### Ethical considerations

Ethical approval for the broader study, titled ‘Determinant of time to presentation for stroke care in Ondo City’, was granted by the Research Ethics Committee of the University of Medical Sciences Teaching Hospital, Ondo State. The committee’s final determination was made on 17 July 2024, and the approval notice was issued on 19 July 2024. The present article uses secondary hospital-record data collected under that approval. During data extraction from hospital records, the authors had access to information that could identify individual participants. However, once extraction was complete, all identifying details were removed, and the resulting analytical dataset was anonymised before analysis. The anonymised dataset was stored securely and used exclusively for the approved research purposes. The findings are presented in aggregate form, ensuring that no information that could identify individual participants is included.

## Results

### Patient characteristics and bivariate associations

Table 1 presents the distribution of early and late presentation by patient characteristics and care-seeking indicators. Late presentation was common across age, sex, marital status, residence and ethnicity categories, and none of these differences reached statistical significance. A statistically significant difference was observed by employment status: 87.9% of employed cases were late, compared with 77.8% of unemployed cases (chi-square = 4.83, p = 0.028).

**Table 1.**
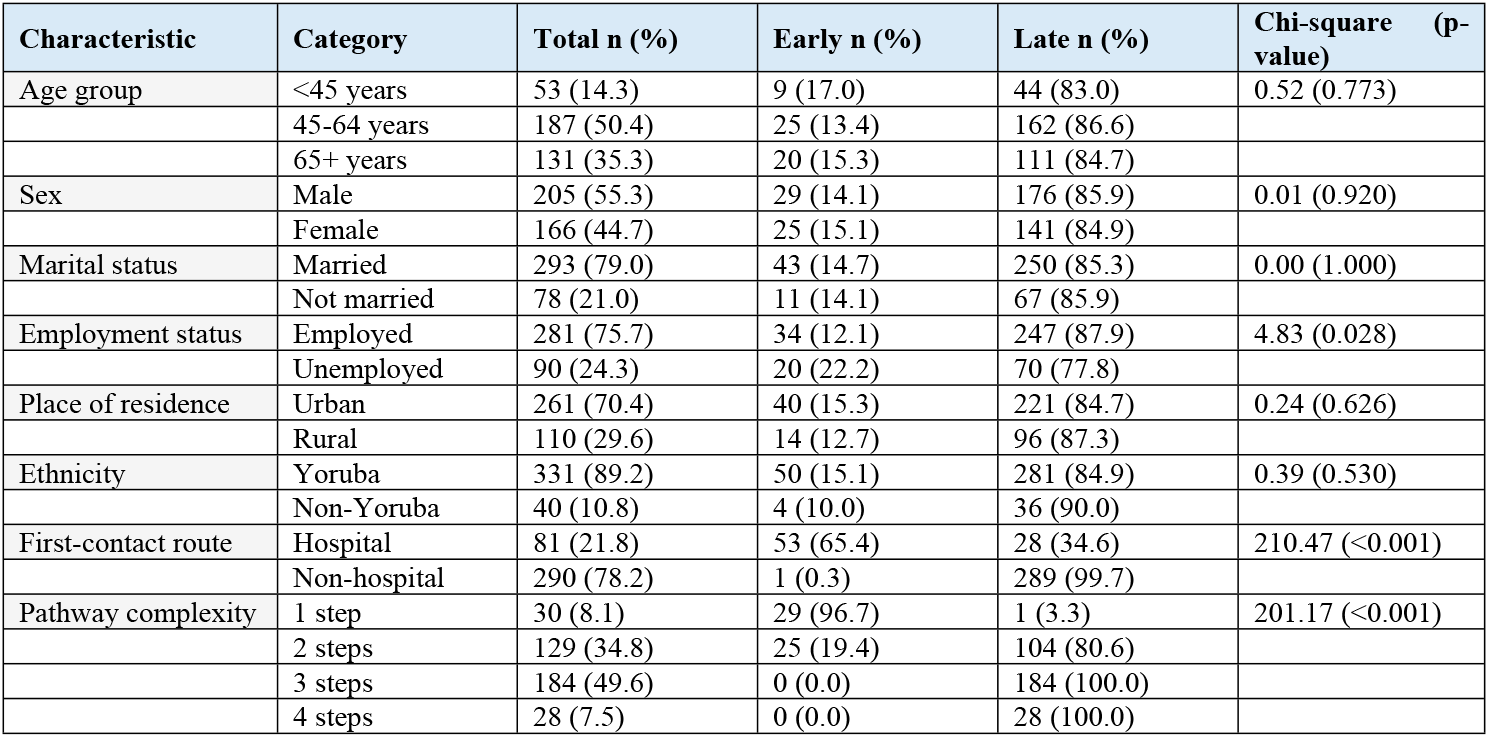
Characteristics of recorded stroke cases by timing of presentation at UNIMEDTH, Ondo State, Nigeria (n = 371)

The clearest crude differences were linked to the care pathway. Among cases with first documented contact in a hospital, 34.6% were late; among cases with first contact outside a hospital, 99.7% were late (chi-square = 210.47, p < 0.001). Pathway complexity showed a corresponding gradient: late presentation occurred in 3.3% of one-step pathways, 80.6% of two-step pathways, and in all recorded three- and four-step pathways (chi-square = 201.17, p < 0.001).

### Prevalence and time to presentation

Table 2 presents the prevalence and time to presentation for stroke care. The analysis included 371 recorded stroke cases.

**Table 2.** Prevalence of late stroke presentation at UNIMEDTH.

| Measures | Overall | Early ($\leq 4$ h) | Late ($> 4$ h) |
| --- | --- | --- | --- |
| Cases, n (%) | 371 (100.0) | 54 (14.6) | 317 (85.4) |
| Median time, h (IQR) | 24 (9-72) | 2 (1-3) | 48 (24-72) |

Fifty-four cases (14.6%) presented within four hours of symptom onset, while 317 (85.4%) were late presentations, arriving after four hours. The median time from symptom onset to presentation was 24 hours (IQR: 9-72 hours). Early presenters had a median presentation time of 2 hours (IQR: 1-3 hours), whereas late presenters presented after a median of 48 hours (IQR: 24-72 hours).

### Adjusted correlates of late presentation

Table 3 presents the modified Poisson regression results. After adjustment for age group, sex, marital status, employment status, residence, ethnicity and first-contact route, non-hospital first contact was the only variable statistically associated with late presentation. Cases with a non-hospital first contact had 2.89 times the prevalence of late presentation compared with cases with a hospital-based first contact (aPR = 2.89, 95% CI: 2.15-3.90; p < 0.001). The estimates for age group, sex, marital status, employment status, residence and ethnicity were not statistically significant after adjustment.

**Table 3.**
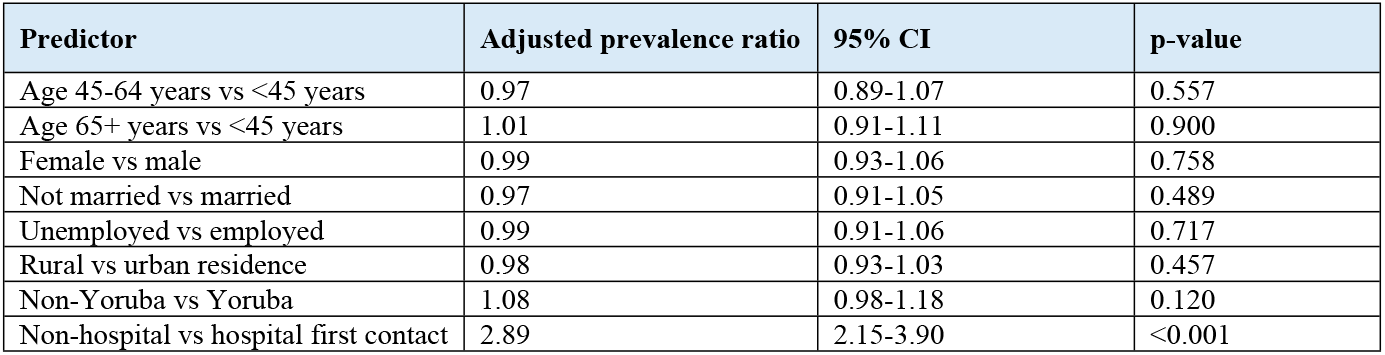
Modified Poisson regression estimates for late presentation for stroke care.

| Predictor | Adjusted prevalence ratio | 95% CI | p-value |
| --- | --- | --- | --- |
| Age 45-64 years vs <45 years | 0.97 | 0.89-1.07 | 0.557 |
| Age 65+ years vs <45 years | 1.01 | 0.91-1.11 | 0.900 |
| Female vs male | 0.99 | 0.93-1.06 | 0.758 |
| Not married vs married | 0.97 | 0.91-1.05 | 0.489 |
| Unemployed vs employed | 0.99 | 0.91-1.06 | 0.717 |
| Rural vs urban residence | 0.98 | 0.93-1.03 | 0.457 |
| Non-Yoruba vs Yoruba | 1.08 | 0.98-1.18 | 0.120 |
| Non-hospital vs hospital first contact | 2.89 | 2.15-3.90 | <0.001 |

The adjusted estimates and their 95% confidence intervals are displayed in Fig 1. The point estimate for non-hospital first contact was markedly elevated, whereas estimates for the sociodemographic variables were clustered near the null value.

**Fig 1.**
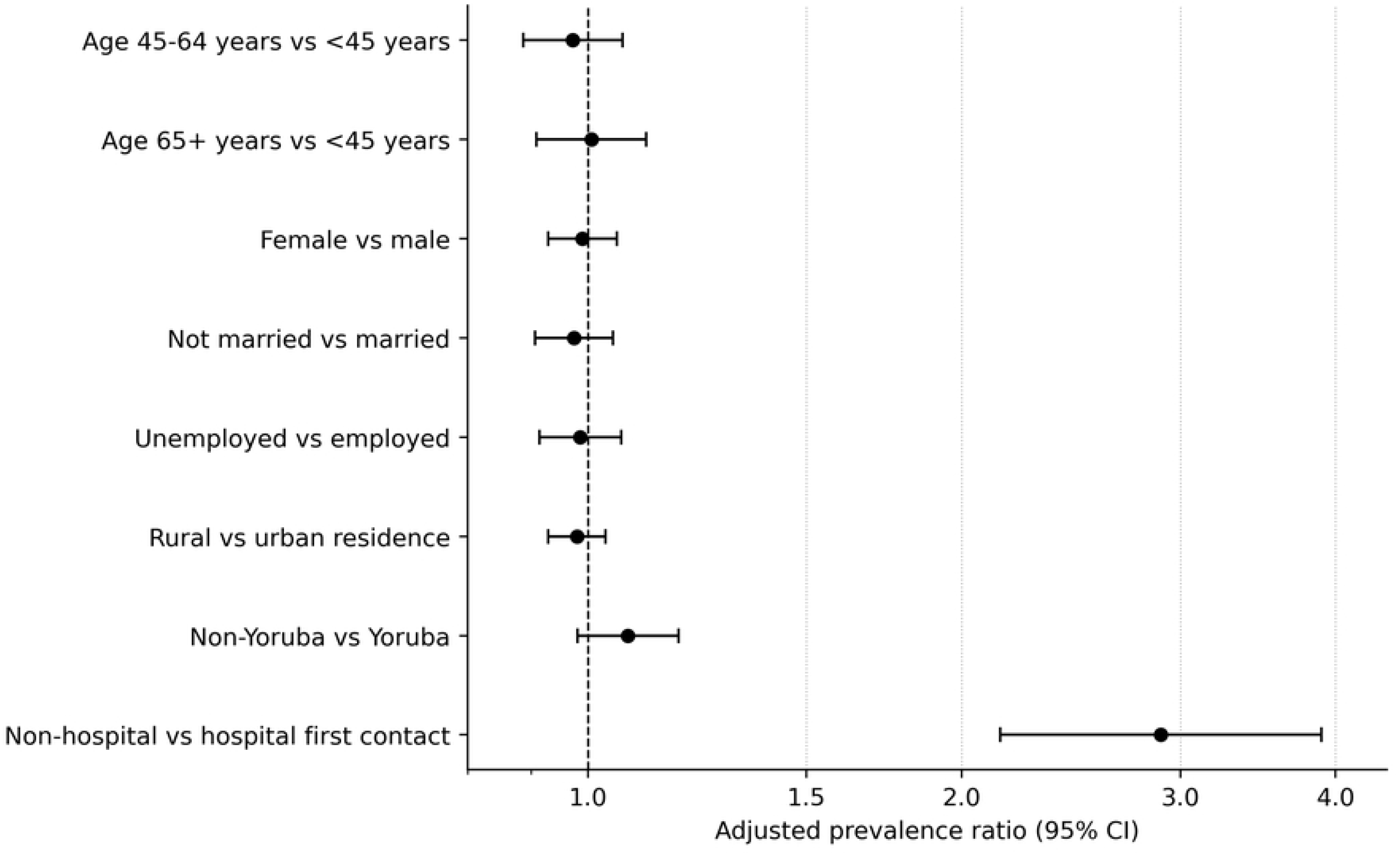
Adjusted prevalence ratios for late presentation among recorded stroke cases at UNIMEDTH.

Table 4 presents the Sensitivity analysis comparing late-presentation classification using >4-hour and >4.5-hour thresholds. Using a late-presentation threshold of more than 4.5 hours yielded the same classification as the primary definition of more than 4 hours: 317 cases were classified as late, and 54 as early under both definitions.

**Table 4.** Sensitivity analysis comparing late-presentation classification using >4-hour and >4.5-hour thresholds.

| Classification threshold | Early presentation<br>n (%) | Late presentation<br>n (%) | Total n (%) | Classification<br>change |
| --- | --- | --- | --- | --- |
| >4 hours after symptom onset | 54 (14.6) | 317 (85.4) | 371 (100.0) | Ref |
| >4.5 hours after symptom onset | 54 (14.6) | 317 (85.4) | 371 (100.0) | No change |

The result occurred because no recorded presentation interval fell between 4.0 and 4.5 hours; therefore, the findings are insensitive to these alternative cut-offs in this dataset.

## Discussion

This study examined late presentation for stroke care using secondary records from a tertiary hospital in Ondo State, Nigeria. Three principal findings emerged. First, late presentation was pervasive: more than four in five recorded cases arrived more than four hours after symptom onset, and the median time to arrival was 24 hours. Second, basic sociodemographic characteristics did not independently distinguish early from late presentation in the adjusted analysis. Third, the first documented care route was strongly associated with delay: cases entering non-hospital routes had a substantially higher prevalence of late presentation than cases whose first contact was hospital-based.

The prevalence of late presentation observed in Ondo State is consistent with African and Nigerian literature. The recent African meta-analysis reported a pooled prehospital delay prevalence of about 80%, while earlier reviews of African stroke care documented substantial gaps in acute stroke system capacity [6,22]. Nigerian studies have likewise reported late hospital or imaging presentation, as evidenced by studies from Calabar and Ibadan [23,24]. Taken together, these findings indicate that late presentation is not an isolated institutional failure; it is a recurring bottleneck in Nigerian and wider African stroke care.

The principal contribution of this study is its focus on the first-contact route. Patients and families may respond to stroke symptoms quickly, yet initially seek help through non-hospital routes that are poorly positioned to provide immediate neuroimaging, thrombolysis eligibility assessment, acute blood-pressure management, complication prevention or urgent referral. In the present records, non-hospital first contacts included home management, use of medicine vendors, traditional remedies, traditional healers, and prayer-based contacts. This should not be interpreted as proving that any single contact type caused delay. It indicates that non-hospital entry into the pathway is a strong marker of late tertiary arrival and a practical intervention point.

This interpretation is consistent with Andersen’s model and with pluralistic accounts of healthcare use in Nigeria. Predisposing and enabling characteristics structure access, but realised access also depends on the pathway households navigate through illness [25,27,32]. Previous Nigerian work shows that formal and informal service use is shaped by context, affordability, proximity and social interpretation of illness [25]. In this stroke cohort, once the first-contact route was taken into account, the recorded background characteristics contributed little independent explanation. For an acute, time-dependent condition, therefore, the first direction of help-seeking appears more consequential than sociodemographic position alone.

The association between increasing pathway complexity and delay reinforces this point. Nearly all one-step pathways were early, while all three- and four-step pathways were late. The result is descriptive rather than causal: multiple pathway steps are likely to be both a consequence and a marker of time already lost after symptom onset. For that reason, pathway complexity was not included in the adjusted model alongside the first-contact route. This modelling decision avoids treating a downstream feature of care navigation as an independent baseline determinant.

The public health implications are immediate. Stroke messages should not stop at naming warning signs; they must specify the required first action: go directly to a hospital or emergency point capable of assessment and referral. This aligns with current stroke-system recommendations that prioritise rapid recognition, emergency response, imaging and organised referral [7,8,19-21]. Because many patients may still first encounter patent medicine vendors, faith-based providers, traditional providers, private clinics or home-care networks, interventions should engage these actors as referral accelerators rather than ignore their role in the pathway. In a context where ambulance systems and stroke units remain limited [9,10], practical gains may come from clearer triage scripts, community referral agreements, transport planning and public messaging that treats stroke as a hospital emergency from the first minute.

This study has limitations. It was based on secondary records from patients who reached a tertiary hospital, and it therefore cannot account for stroke cases managed elsewhere or cases that never reached hospital care. The completeness and precision of onset times and prior care contacts depend on documentation within the record sources. The dataset did not consistently record information on stroke subtype, stroke severity, educational attainment, household resources, symptom recognition, transport constraints, or distance to hospital; these factors could influence presentation time. In addition, the first-contact comparison included only one early case in the non-hospital category, indicating sparse data and requiring caution about the magnitude of the adjusted association. Finally, the retrospective observational design supports statements about association, not causal effects.

## Conclusion

Late presentation for stroke care was highly prevalent among stroke cases identified from UNIMEDTH records in Ondo State. While delay occurred across the sociodemographic categories available in the data, non-hospital first contact was strongly associated with arrival after the early treatment window. Strategies to improve timely stroke presentation should therefore prioritise rapid hospital-directed responses to suspected stroke symptoms and strengthen urgent referral from common non-hospital first-contact points. Further prospective research should examine how symptom recognition, household decision-making, transport and stroke severity interact with care pathways to produce late presentation.

## Data Availability

The data underpinning this study's findings are drawn from retrospective hospital records, including the Stroke Registry of the University of Medical Sciences Teaching Hospital, radiology department records, referral notes, and ambulance records. The dataset contains sensitive patient-level clinical and care-seeking information. It cannot be made publicly available, as the ethics approval did not cover public deposition and could compromise participant confidentiality. Access to the anonymised dataset may be granted to qualified researchers upon reasonable request and subject to approval by the Research Ethics Committee of the University of Medical Sciences Teaching Hospital, Ondo State, Nigeria. Requests for data access should be directed to the Research Ethics Committee at the University of Medical Sciences Teaching Hospital, Ondo State, Nigeria.

## References

1. Feigin VL, Brainin M, Norrving B, Martins SO, Pandian J, Lindsay P, et al. World Stroke Organisation: Global Stroke Fact Sheet 2025. Int J Stroke. 2025;20(2):132–144. doi:10.1177/17474930241308142.

2. GBD 2021 Stroke Risk Factor Collaborators. Global, regional, and national burden of stroke and its risk factors, 1990-2021: a systematic analysis for the Global Burden of Disease Study 2021. Lancet Neurol. 2024;23(10):973–1003. doi:10.1016/S1474-4422(24)00369-7.

3. Prust ML, Forman R, Ovbiagele B. Addressing disparities in the global epidemiology of stroke. Nat Rev Neurol. 2024;20(4):207–221. doi:10.1038/s41582-023-00921-z.

4. Okekunle AP, Jones S, Adeniji O, Watkins C, Hackett M, Di Tanna GL, et al. Stroke in Africa: a systematic review and meta-analysis of the incidence and case-fatality rates. Int J Stroke. 2023;18(6):634–644. doi:10.1177/17474930221147164.

5. Adoukonou T, Kossi O, Fotso Mefo P, Agbetou M, Magne J, Gbaguidi G, et al. Stroke case fatality in sub-Saharan Africa: systematic review and meta-analysis. Int J Stroke. 2021;16(8):902–916. doi:10.1177/1747493021990945.

6. Urimubenshi G, Cadilhac DA, Kagwiza JN, Wu O, Langhorne P. Stroke care in Africa: a systematic review of the literature. Int J Stroke. 2018;13(8):797–805. doi:10.1177/1747493018772747.

7. Pandian JD, Kalkonde Y, Sebastian IA, Felix C, Urimubenshi G, Bosch J. Stroke systems of care in low-income and middle-income countries: challenges and opportunities. Lancet. 2020;396(10260):1443–1451. doi:10.1016/S0140-6736(20)31374-X.

8. Bosch J, Lotlikar R, Melifonwu R, Roushdy T, Sebastian IA, Abraham SV, et al. Prehospital stroke care in low- and middle-income countries: a World Stroke Organisation Scientific Statement. Int J Stroke. 2025;20(8):918–927. doi:10.1177/17474930251351867.

9. Arabambi B, Oshinaike O, Ogun SA, Eze C, Bello AH, Igetei S, et al. Stroke units in Nigeria: a report from a nationwide organisational cross-sectional survey. Pan Afr Med J. 2022;42:140. doi:10.11604/pamj.2022.42.140.35086.

10. Oyedokun TO, Islam EM, Eke NO, Oladipo O, Akinola OO, Salami O. Out-of-hospital emergency care in Nigeria: a narrative review. Afr J Emerg Med. 2023;13(3):171–176. doi:10.1016/j.afjem.2023.06.001.

11. The National Institute of Neurological Disorders and Stroke rt-PA Stroke Study Group. Tissue plasminogen activator for acute ischemic stroke. N Engl J Med. 1995;333(24):1581–1587. doi:10.1056/NEJM199512143332401.

12. Hacke W, Kaste M, Bluhmki E, Brozman M, Davalos A, Guidetti D, et al. Thrombolysis with alteplase 3 to 4.5 hours after an acute ischemic stroke. N Engl J Med. 2008;359(13):1317–1329. doi:10.1056/NEJMoa0804656.

13. Emberson J, Lees KR, Lyden P, Blackwell L, Albers G, Bluhmki E, Cohen G, et al. Effect of treatment delay, age, and stroke severity on the effects of intravenous thrombolysis with alteplase for acute ischaemic stroke: a meta-analysis of individual patient data from randomised trials. Lancet. 2014;384(9958):1929–1935. doi:10.1016/S0140-6736(14)60584-5.

14. Saver JL. Time is brain--quantified. Stroke. 2006;37(1):263–266. doi:10.1161/01.STR.0000196957.55928.ab.

15. Goyal M, Menon BK, van Zwam WH, Dippel DWJ, Mitchell PJ, Demchuk AM, et al. Endovascular thrombectomy after large-vessel ischaemic stroke: a meta-analysis of individual patient data from five randomised trials. Lancet. 2016;387(10029):1723–1731. doi:10.1016/S0140-6736(16)00163-X.

16. Nogueira RG, Jadhav AP, Haussen DC, Bonafe A, Budzik RF, Bhuva P, et al. Thrombectomy 6 to 24 hours after stroke with a mismatch between deficit and infarct. N Engl J Med. 2018;378(1):11–21. doi:10.1056/NEJMoa1706442.

17. Albers GW, Marks MP, Kemp S, Christensen S, Tsai JP, Ortega-Gutierrez S, et al. Thrombectomy for stroke at 6 to 16 hours with selection by perfusion imaging. N Engl J Med. 2018;378(8):708–718. doi:10.1056/NEJMoa1713973.

18. Ma H, Campbell BCV, Parsons MW, Churilov L, Levi CR, Hsu C, et al. Thrombolysis guided by perfusion imaging up to 9 hours after the onset of stroke. N Engl J Med. 2019;380(19):1795–1803. doi:10.1056/NEJMoa1813046.

19. Powers WJ, Rabinstein AA, Ackerson T, Adeoye OM, Bambakidis NC, Becker K, et al. Guidelines for the early management of patients with acute ischemic stroke: 2019 update to the 2018 guidelines for the early management of acute ischemic stroke. Stroke. 2019;50(12):e344–e418. doi:10.1161/STR.0000000000000211.

20. Prabhakaran S, Gonzalez NR, Zachrison KS, Adeoye O, Alexandrov AW, Ansari SA, et al. 2026 Guideline for the early management of patients with acute ischemic stroke: a guideline from the American Heart Association/American Stroke Association. Stroke. 2026. doi:10.1161/STR.0000000000000513.

21. Berge E, Whiteley W, Audebert H, De Marchis GM, Fonseca AC, Padiglioni C, et al. European Stroke Organisation guidelines on intravenous thrombolysis for acute ischaemic stroke. Eur Stroke J. 2021;6(1): I–LXII. doi:10.1177/2396987321989865.

22. Ganeti DD, Dulo AO, Ilala BW, Benti NB, Diriba M, Abdulwehab S, et al. Prehospital delay and associated factors among stroke patients in Africa: a systematic review and meta-analysis. PLoS One. 2025;20(6):e0326323. doi:10.1371/journal.pone.0326323.

23. Philip-Ephraim EE, Charidimou A, Otu AA, Eyong EK, Williams UE, Ephraim RP. Factors associated with prehospital delay among stroke patients in a developing African country. Int J Stroke. 2015;10(4):E39. doi:10.1111/ijs.12469.

24. Ogbole GI, Owolabi MO, Ogun O, Ogunseyinde OA, Ogunniyi A. Time of presentation of stroke patients for CT imaging in a Nigerian tertiary hospital. Ann Ib Postgrad Med. 2015;13(1):23–28.

25. Fayehun O, Ajisola M, Uthman O, Oyebode O, Oladejo A, Owoaje E, et al. A contextual exploration of healthcare service use in urban slums in Nigeria. PLoS One. 2022;17(2):e0264725. doi:10.1371/journal.pone.0264725.

26. Kleinman A. Patients and healers in the context of culture: an exploration of the borderland between anthropology, medicine, and psychiatry. Berkeley: University of California Press; 1980.

27. Kroeger A. Anthropological and socio-medical health care research in developing countries. Soc Sci Med. 1983;17(3):147–161. doi:10.1016/0277-9536(83)90248-4.

28. Obembe AO, Olaogun MO, Bamikole AA, Komolafe MA, Odetunde MO. Awareness of risk factors and warning signs of stroke in a Nigerian university. J Stroke Cerebrovasc Dis. 2014;23(4):749–758. doi:10.1016/j.jstrokecerebrovasdis.2013.06.036.

29. Wahab KW, Okokhere PO, Ugheoke AJ, Oziegbe O, Asalu AF, Salami TA. Awareness of warning signs among suburban Nigerians at high risk for stroke is poor: a cross-sectional study. BMC Neurol. 2008;8:18. doi:10.1186/1471-2377-8-18.

30. Wahab KW, Kayode OO, Musa OI. Knowledge of stroke risk factors among Nigerians at high risk. J Stroke Cerebrovasc Dis. 2015;24(1):125–129. doi:10.1016/j.jstrokecerebrovasdis.2014.07.053.

31. Kayode-Iyasere E, Odiase FE. Awareness of stroke, its warning signs, and risk factors in the community: a study from the urban population of Benin City, Nigeria. Sahel Med J. 2019;22(3):134–139. doi:10.4103/smj.smj_4_18.

32. Andersen RM. Revisiting the behavioural model and access to medical care: does it matter? J Health Soc Behav. 1995;36(1):1–10. doi:10.2307/2137284.

33. Zou G. A modified Poisson regression approach to prospective studies with binary data. Am J Epidemiol. 2004;159(7):702–706. doi:10.1093/aje/kwh090.

34. Chen W, Qian L, Shi J, Franklin M. Comparing performance between log-binomial and robust Poisson regression models for estimating risk ratios under model misspecification. BMC Med Res Methodol. 2018;18:63. doi:10.1186/s12874-018-0519-5.

